# The impact of changes in diagnostic testing practices on estimates of COVID-19 transmission in the United States

**DOI:** 10.1101/2020.04.20.20073338

**Authors:** Virginia E. Pitzer, Melanie Chitwood, Joshua Havumaki, Nicolas A. Menzies, Stephanie Perniciaro, Joshua L. Warren, Daniel M. Weinberger, Ted Cohen

## Abstract

Estimates of the reproductive number for novel pathogens such as SARS-CoV-2 are essential for understanding the potential trajectory of the epidemic and the level of intervention that is needed to bring the epidemic under control. However, most methods for estimating the basic reproductive number (*R*_0_) and time-varying effective reproductive number (*R*_*t*_) assume that the fraction of cases detected and reported is constant through time. We explore the impact of secular changes in diagnostic testing and reporting on estimates of *R*_0_ and *R*_*t*_ using simulated data. We then compare these patterns to data on reported cases of COVID-19 and testing practices from different United States (US) states. We find that changes in testing practices and delays in reporting can result in biased estimates of *R*_0_ and *R*_*t*_. Examination of changes in the daily number of tests conducted and the percent of patients testing positive may be helpful for identifying the potential direction of bias. Changes in diagnostic testing and reporting processes should be monitored and taken into consideration when interpreting estimates of the reproductive number of COVID-19.

## Introduction

The initial stages of the COVID-19 epidemic in the United States (US) were characterized by difficulties in delivering and administering diagnostic tests (1). First, the real-time quantitative PCR assay developed and distributed by the US Centers for Disease Control and Prevention (CDC) suffered from performance issues (2, 3). As a result, all initial and confirmatory testing needed to be carried out by the CDC, which led to reporting delays and capacity issues early in the epidemic (4, 5). Initially, tests were only administered to individuals with a history of travel to certain countries or known contact with a positive case. By the time testing capacity increased, state and local health departments were faced with heavy demand for COVID-19 testing. Only individuals meeting specific criteria could receive a test, and these criteria have varied from state to state and over time (see Supporting Information (SI) Dataset).

Concurrently, mathematical modelers have been analyzing data on reported COVID-19 cases in order to develop forecasts of future incidence and evaluate the potential impact of social distancing and other control measures, often at the behest of policymakers and public health officials. These models typically rely on estimates of the reproductive number of the virus. The basic reproductive number (*R*_0_) is defined as the expected number of secondary infections produced by an infectious individual in a fully susceptible population; this can be used to derive the expected fraction of the population that will become infected in the absence of interventions and the level of control and/or immunity that is needed to eliminate the pathogen from circulation (6). The time-varying effective reproductive number (*R*_*t*_) measures the average number of secondary infections per case at each time-point in the epidemic (6), and can be used for real-time monitoring of the impact of control measures (7–9). As control measures are implemented and immunity increases in the population, *R*_*t*_ will decrease (6, 10). The value of *R*_0_ can be estimated from the growth rate of the number of cases early on in the epidemic and estimates of the distribution of the generation time (i.e. the time between infection events of successive cases in a transmission chain) (11), whereas instantaneous values of *R*_*t*_ can be estimated based on the time series of case notifications and the distribution of the generation time (7). These methods have been shown to be robust to under-detection and underreporting, so long as the probability that a true case is detected and reported remains constant through time.

Here, we use simulations to explore the potential magnitude and direction of biases introduced by changes in diagnostic testing and reporting practices similar to those occurring during the early stages of the COVID-19 epidemic in the US. We then compute preliminary estimates of *R*_0_ and *R*_*t*_ for different states, based on publicly available data from The COVID Track Project (12). We examine changes in testing practices and trends of the cumulative number of tests reported to evaluate the potential that these estimates are biased.

## Results

Based on our simulations (Table S1, Fig. S1), the likelihood and degree to which *R*_0_ and *R*_*t*_ are biased depends on the manner in which diagnostic testing practices and reporting changes over time. When the fraction of incident cases detected and reported is constant over time and testing capacity scales with the number of “true” cases, the number of confirmed positive cases provides an unbiased estimate of *R*_0_, despite possible delays in the reporting process (Fig. 1A, Fig. S2). In this instance, the percent of individuals testing positive is expected to be stable over time. Estimates of *R*_*t*_ are also expected to be unbiased after the first 5-6 days (approximately equal to the mean reporting delay), but lag 2-4 days behind in detecting a decrease in *R*_*t*_ below the threshold value of 1 (i.e. when the epidemic is receding). If the fraction of true cases detected and reported is increasing or decreasing linearly over time, estimates of *R*_0_ based on the growth rate of confirmed cases will be over- or under-estimates, respectively, of the true *R*_0_ (Fig. 1B-C). The time-varying reproductive number, *R*_*t*_, will also be slightly over- or underestimated, especially early on when there is a reporting delay. A gradual increase or decrease in the percent of individuals testing positive is a potential indicator of such bias. However, the percent positive is also expected to decrease or increase over time if the testing capacity expands more or less quickly than the number of true cases, respectively (Fig. 1D-E). In this instance, estimates of *R*_0_ and *R*_*t*_ based on the number of confirmed cases are unbiased (Fig. S2).

**Figure 1.**
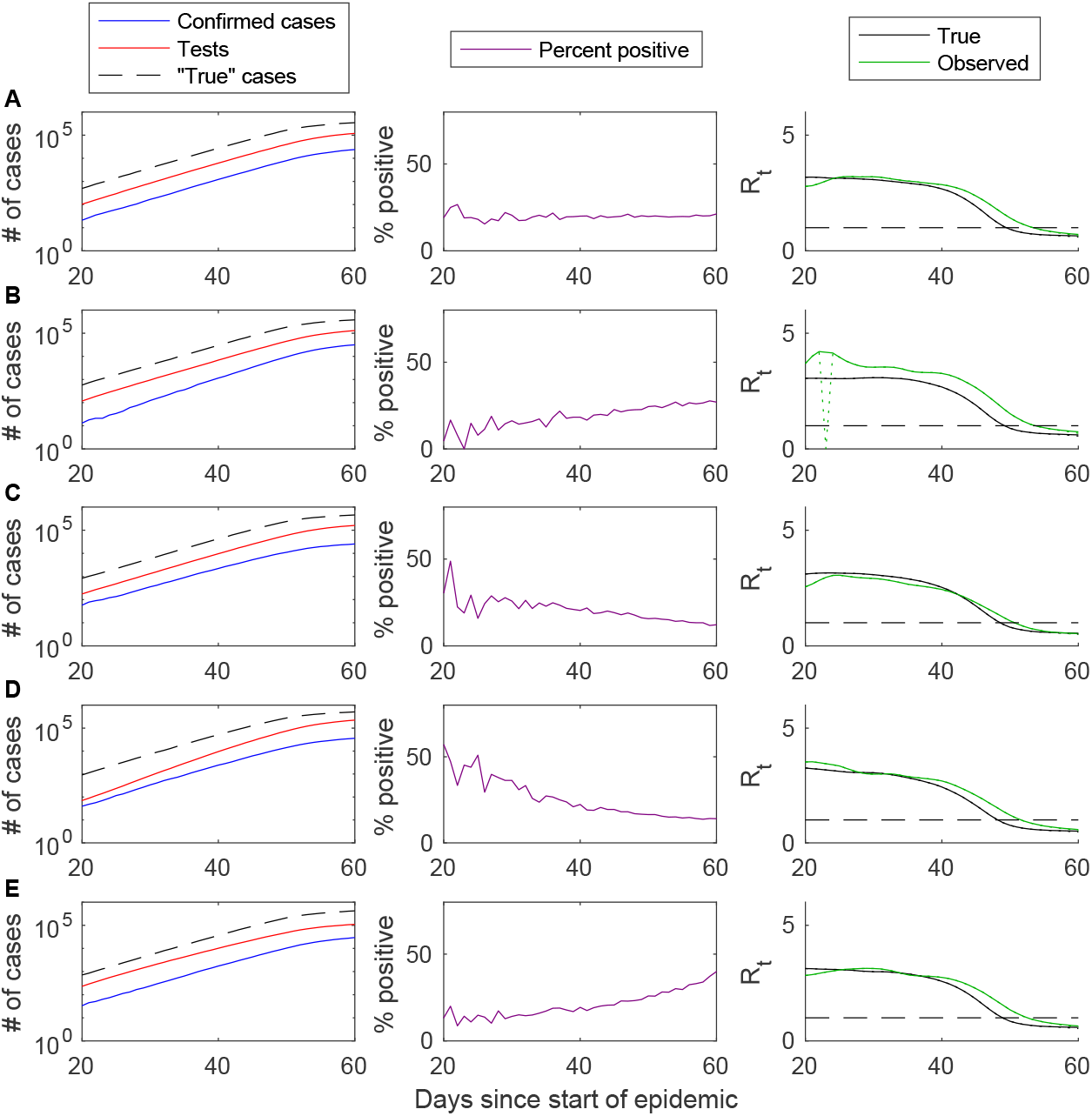
Impact of gradual changes in testing practices on *R*_t_ estimation based on simulated data. The cumulative number of confirmed cases (blue), individuals tested (red), and “true” infections (dashed black line) are plotted on the log_10_ scale for days 20 to 60 of the simulated epidemic (left), along with the percent of tests positive (purple, middle) and the estimated time-varying reproductive number (*R*_*t*_, right) for the true cases (black) and confirmed cases (green). (A) Base case in which the fraction of true cases detected and reported and the reporting delay are constant over time and the testing capacity scales with the number of true cases. (B) The fraction of true cases detected increases from 5% to 15% between days 20 and 60. (C) The fraction of cases detected decreases from 15% to 5% between days 20 and 60. (D) The testing capacity increases from 0.2 individuals per case to 0.8 individuals per case between days 20 and 60. (E) The testing capacity decreases from 0.8 individuals per case to 0.2 individuals per case between days 20 and 60.

Abrupt changes to testing criteria, affecting the fraction of true cases detected and reported, are also expected to bias estimates of *R*_0_ and lead to a large (up to two-fold) but temporary bias in estimates of *R*_*t*_ (Fig. 2A-B). The potential for such bias may be indicated by a sudden change in the percent of individuals testing positive, as well as a temporary change to the slope of the log of cumulative cases. A similar change in the percent positive may also occur with an abrupt change to the testing capacity (Fig. 2C-D). However, in this case, it is accompanied by a change to the slope of the log of cumulative individuals tested, and estimates of *R*_0_ and *R*_*t*_ based on fitting to the number of positive cases are not expected to be biased. The most difficult bias to detect may be due to a change in the reporting delay distribution (Fig. 2E-F). In this case, the percent positive is likely to remain roughly constant through time, but estimates of *R*_0_ and *R*_*t*_ will be biased, especially when the reporting delay increases.

**Figure 2.**
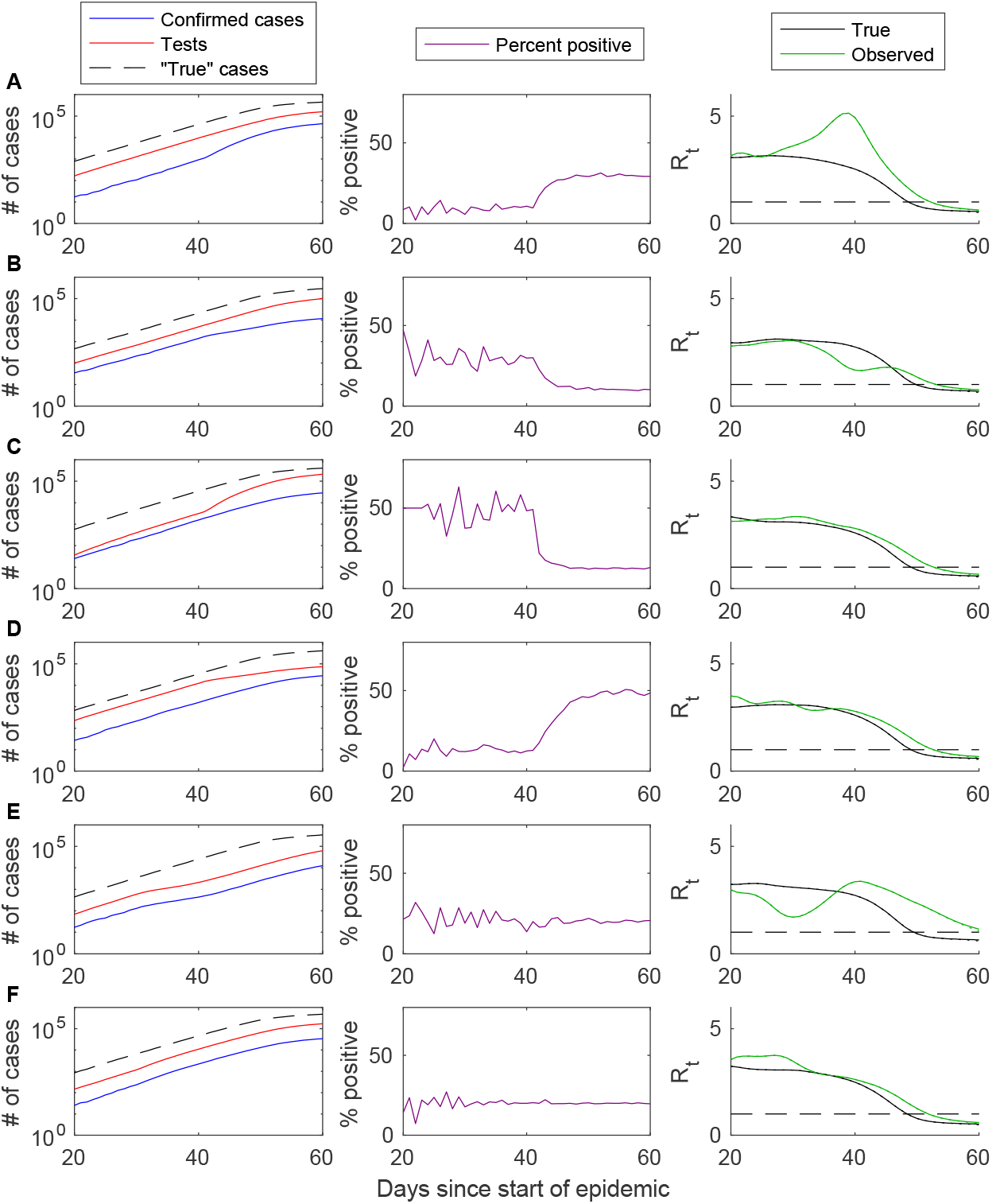
Impact of abrupt changes in testing practices on *R*_t_ estimation based on simulated data. The cumulative number of confirmed cases (blue), individuals tested (red), and “true” infections (dashed black line) are plotted on the log_10_ scale for days 20 to 60 of the simulated epidemic (left), along with the percent of tests positive (purple, middle) and the estimated time-varying reproductive number (*R*_*t*_, right) for the true cases (black) and confirmed cases (green). (A) The fraction of true cases detected and reported increases from 5% to 15% on day 40. (B) The fraction of true cases detected and reported decreases from 15% to 5% on day 40. (C) The testing capacity increases from 0.2 individuals per case to 0.8 individuals per case on day 40. (D) The testing ratio decreases from 0.8 individuals per case to 0.2 individuals per case on day 40. (E) The mean reporting delay increases from 6.6 days to 13.2 days on day 40. (F) The mean reporting delay decreases from 6.6 days to 3.3 days on day 40.

When the bias is due to a change in the testing criteria, affecting the fraction of true cases detected and reported, fitting models to the total number of individuals tested over time can recover unbiased estimates of the reproductive numbers (Fig. S2-S3). Thus, when the percent of positive tests is changing through time and there is uncertainty over whether it is due to changes in the testing probability or the testing ratio, fitting models to both the number of positive cases and the total number of tests may provide bounds on the uncertainty in estimates of *R*_0_ and *R*_*t*_. However, this approach cannot correct for bias due to changes in the reporting delay distribution.

Based on the number of confirmed COVID-19 cases across the US through March 24, 2020 (before any observable impact of social distancing measures, Fig. 3), and assuming a fixed serial interval of 6.5 days, we estimate that *R*_0_ for the US is 3.45 (95% confidence interval (CI): 3.44-3.46, accounting *only* for uncertainty in the growth rate). Estimates of *R*_0_ for all 50 states and the District of Columbia vary from 1.92 (95% CI: 1.61-2.27, South Dakota) to 5.17 (95% CI: 4.75-5.65, Missouri) (Table S2). Estimates of *R*_0_ based on the growth rate of the number of tests performed are similar for the entire US, but are slightly larger on average for the individual states (Table S2). Nationally, the percent of individuals testing positive for COVID-19 decreased from 20-25% in early March to around 15% in mid-March. This early decline in the percent positive is likely attributable to an increase in testing capacity, rather than a decrease in the fraction of true cases detected and reported; therefore, estimates of *R*_0_ based on the number of confirmed cases should be unbiased. However, trends in the percent of tests positive vary by state (Fig. 4).

**Figure 3.**
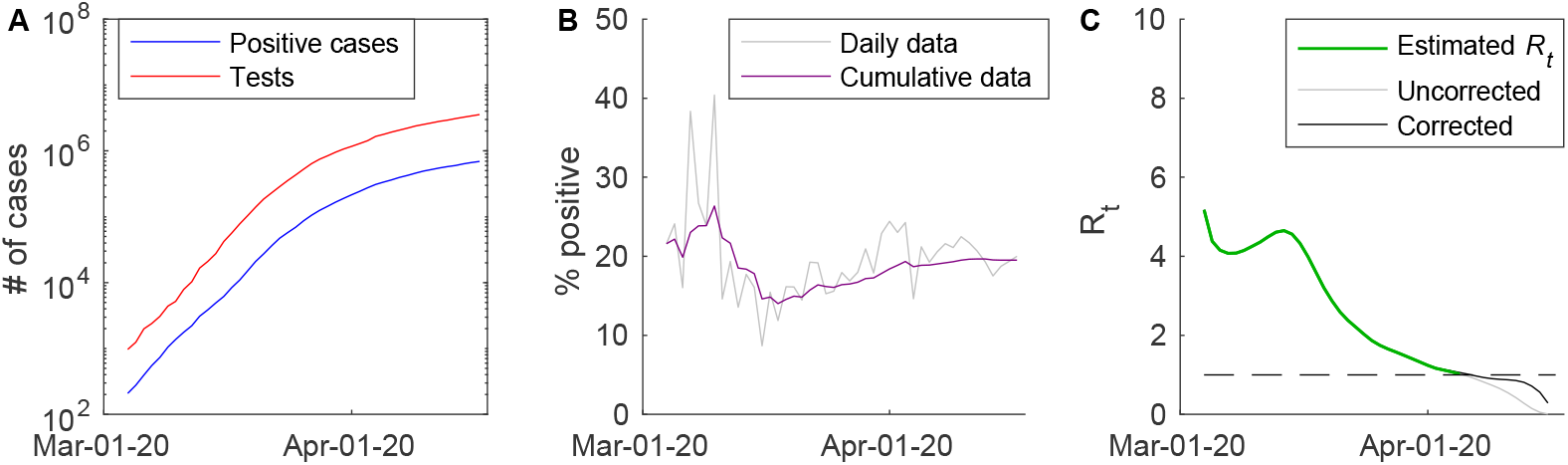
Reported number of COVID-19 cases and tests in the US and estimated time-varying reproductive number. (A) The cumulative number of confirmed cases (blue) and individuals tested (red) are plotted on the log_10_ scale for March 4 to April 17, 2020. (B) The percent of tests positive through time is plotted for the daily data (grey) and the cumulative data (purple). (C) The estimated value of the time-varying reproductive number, *R*_*t*_, is plotted for March 4 to April 5 (green), which is the last day that the value of *R*_*t*_ can be reliably estimated. Uncorrected (grey) and corrected (black) values of *R*_*t*_ are plotted for April 2 to April 16.

**Figure 4.**
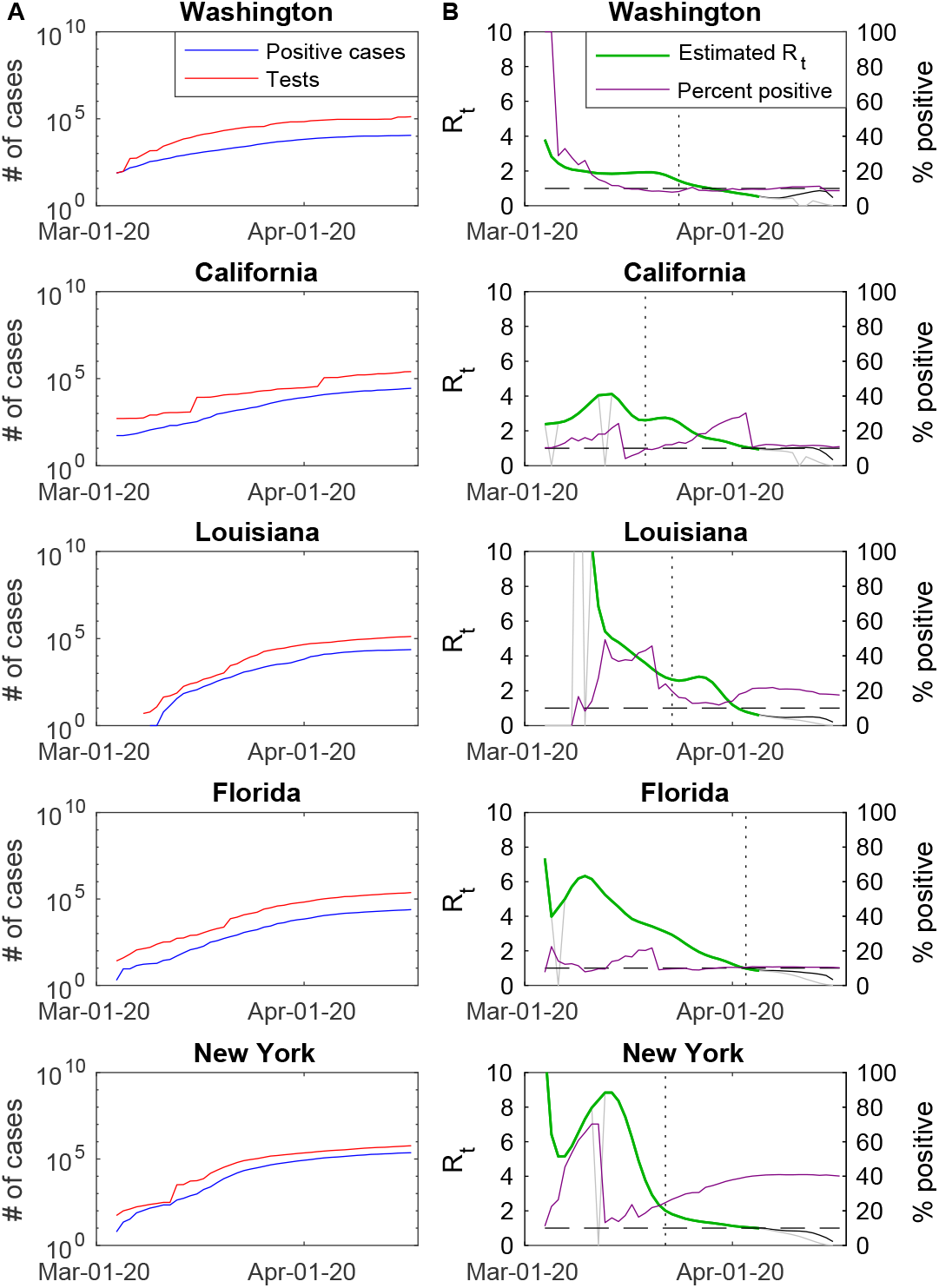
Reported number of COVID-19 cases and tests and estimated time-varying reproductive numbers for select US states. (A) The cumulative number of confirmed cases (blue) and individuals tested (red) are plotted on the log_10_ scale for March 4 to April 17, 2020. (B) The percent of tests positive (purple) and the estimated value of the time-varying reproductive number (*R*_*t*_, green) are plotted for March 4 to April 17, 2020. April 5 is the last day that the value of *R*_*t*_ can be reliably estimated; uncorrected (grey) and corrected (black) values of *R*_*t*_ are plotted for April 2 to April 16. The vertical dotted lines represent the dates that stay-at-home orders were issued in each state.

Estimates of *R*_*t*_ for the US and individual states were generally high (*R*_*t*_>4) initially but decreased over time (Fig. S4). This may be due to a low probability of detecting cases at the start of the epidemic in late February/early March. In Washington and California, where COVID-19 cases in the US were first recognized, estimates of *R*_*t*_ for early March were slightly lower (Fig. 4). Nationally, *R*_*t*_ had decreased to less than 2 by March 24, 2020. In some states, *R*_*t*_ had already declined substantially before stay-at-home orders were issued. As of April 5, 2020, which is the last reliable day for which we could generate estimates, *R*_*t*_ was less than 1 (meaning the epidemic was starting to decline) in 24 of 51 states, but was hovering around 1 in a number of states (e.g. California) (Fig. 4, Fig. S4).

Nationally (and in most states), the percent of individuals testing positive for COVID-19 has been gradually increasing since mid-March (Fig. 3, Fig. 4). This could be occurring either because the fraction of individuals infected with SARS-CoV-2 detected and reported is increasing, or because states are reaching their testing capacity and individuals more likely to be infected are being prioritized for testing. If it is the former, our analysis suggests that the more recent estimates of *R*_*t*_ may be upwardly biased. Moreover, temporary increases in *R*_*t*_ around mid-March in states like New York and California might be due to early restrictions to testing criteria. However, abrupt decreases in the percent of tests positive were generally due to large increases in the total number of tests reported, and therefore should not reflect a bias in estimates of *R*_*t*_ based on the number of confirmed cases.

## Discussion

Over the first month and a half of the COVID-19 epidemic in the US, testing practices have varied dramatically over time and from state to state (SI Dataset) (13). As of April 17, 2020, the total number of tests reported per capita has varied from 5.7 tests per 1,000 people in Virginia to 29.5 tests per 1,000 people in New York (12). Due to the limited availability of tests early on, most states recommended that only those with a history of travel to affected countries or known contact with a confirmed case be tested (4, 5). Since the disease has become more widespread throughout the US and testing capacity has increased, testing guidelines have been relaxed, but there are still considerable differences from state to state (SI Dataset). For example, as of April 6, 2020, Washington state had no restrictions on who can be tested for COVID-19, but prioritized hospitalized individuals and essential service providers exhibiting symptoms (14). As of April 15, New York still recommended restricting testing to those with a known positive contact or travel history, as well as symptomatic individuals who had tested negative for other infections (15). As testing practices change over time, we have demonstrated that these changes may introduce bias into estimates of *R*_0_ and *R*_*t*_, affecting inference about how much control is needed and when control measures have reduced transmission below the critical threshold necessary to sustain the epidemic.

Our estimate of the mean *R*_0_ in the US of 3.45 is reasonably consistent with published estimates. Early analyses from China report a range for *R*_0_ of 2.24-3.58, assuming a mean serial interval of 8 days and a 2- to 8-fold increase in the reporting rate (16). However, more recent analyses estimate a median *R*_0_ of 5.7 (95% CI 3.8, 8.9) (17). Analysis of data from Europe and the US present a similarly high *R*_0_, with median values ranging from 4.0 to 7.1 when assuming a serial interval of 6-9 days (18). Our estimated mean *R*_0_ is notably lower, but this was based on a similar estimated growth rate of 0.275, suggesting the difference can be attributed to different assumptions about the serial interval. Estimates of *R*_*t*_ have not been widely reported for the US, but other studies have found that *R*_*t*_ decreased from 2-5 at the start of the epidemic to ∼1 following travel restrictions in Wuhan and various European countries (19, 20), which is in line with our findings.

Monitoring the number of tests performed and the percent of tests that are positive over time can help to indicate the potential for bias in estimates of reproductive numbers. However, the reporting of test results, particularly for negative tests, has been inconsistent in many states. In California, for example, there was a more than eight-fold increase in the number of negative tests reported on March 13, and another four-fold increase on April 4, 2020 (12). These large increases in the number of negative tests were not accompanied by a corresponding increase in the number of confirmed cases. Thus, while estimates of *R*_0_ and *R*_*t*_ based on the number of confirmed cases are not expected to be influenced by these abrupt changes in the number of reported tests, it becomes difficult to interpret the intervening gradual increases in the percent of tests positive.

It is more difficult to detect whether the time between onset of infectiousness and the reporting of test results (i.e., the reporting delay) has changed over time. Our simulations suggest that such changes could bias estimates of *R*_0_ and *R*_*t*_, but would not be reflected in the percent of individuals testing positive over time. Individual-level data on the date of symptom onset, date of testing, and date of reporting are needed to resolve this potential bias. Estimates of *R*_*t*_ are also expected to lag behind true changes in the transmission rate due to reporting delays. Now-casting approaches may be useful for resolving this by inferring the number of infections occurring on each day based on the observed cases, hospitalizations and deaths, and known reporting delays (21).

Imminent decisions regarding the lifting of stay-at-home orders and loosening social distancing requirements, and when such measures may need to be reinstated, depend on having a good understanding of current levels of transmission. Reliable estimates of the reproductive number are essential for quantifying the impact of control measures on transmission and making informed decisions about future interventions, e.g. (7, 19, 20, 22, 23). However, changes in testing policies and practices, as well as delays in the reporting process, can lead to bias in estimates of the reproductive number, as we have demonstrated. It important to carefully document and track such changes in testing and reporting practices in order to make correct inferences.

## Methods

### Examining the impact of changes in testing using simulated data

We simulated a stochastic SEIR (Susceptible-Exposed-Infected-Recovered) model to explore the potential impact of changes in testing practices on estimates of *R*_0_ and *R*_*t*_. We modeled a population of 1 million individuals and initialized the epidemic with 10 infectious individuals to minimize the chances of early epidemic fadeout. We assumed everyone else was susceptible at the start of the epidemic. New infections were assumed to arise according to a Poisson process (approximate method); the state transitions and rates are described in Table 1, and model parameters are given in Table 2. We simulated the model to day 70 using a time-step of Δ*t*=0.05 days, and assumed a decrease in the transmission rate occurring on day 50, consistent with the impact of social distancing interventions. Models were run using MATLAB v9.3 (MathWorks, Natick, MA); code is available from https://github.com/vepitzer/COVIDtestingbias.

**Table 1.** State transitions and rates for the stochastic simulation model. Model parameters are defined in Table 2.

| Event | State transition | Rate |
| --- | --- | --- |
| New infection | $S \rightarrow E$ | $\beta I(t-1) \frac{S(t-1)}{N(t-1)} \Delta t$ |
| Onset of infectiousness | $E \rightarrow I$ | $\nu E(t-1) \Delta t$ |
| Recovery from infectiousness | $I \rightarrow R$ | $\gamma I(t-1) \Delta t$ |

**Table 2.** Stochastic SEIR model parameters.

| Parameter | Symbol | Value | Reference |
| --- | --- | --- | --- |
| Transmission parameter | $\beta$ | 0.85 for $d < 50$<br>0.3 for $d \geq 50$ | Assumption<br>(consistent with $R_0 \sim 3$<br>and $R_t \sim 1$) |
| Average duration of latent period | $1/\nu$ | 3.7 days | (24) |
| Average duration of infectious period | $1/\gamma$ | 3.5 days | (24) |

We tracked the number of “true cases” on day *d* (*Y*_*d*_) as the number of individuals entering the infectious period each day. We assumed that each true case occurring on day *d* had a probability *p*_test_(*d*) of being tested, and that for every true case that occurred on day *d*, there were *n*_test_(*d*) individuals with similar symptoms who were tested. Furthermore, we assumed that testing and reporting of positive tests occurred with some delay *ρ*_*d*_(*t*), which followed a gamma distribution with parameters *a*_*d*_ and *b*_*d*_. Thus, we calculated the number of individuals tested (*T*_*d*_) and the number of positive cases (*C*_*d*_) on day *d* as follows:

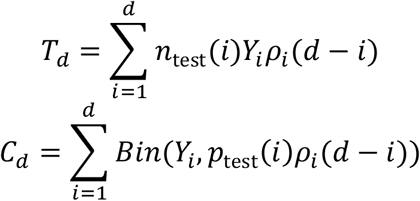

We rounded the value of *T*_*d*_ to the nearest integer and sampled *C*_*d*_ as binomial random variable. We allowed for the observation of cases occurring up to day 95 (even though no new cases were modeled after day 70) to examine the impact of the reporting delay on estimates of *R*_*t*_.

As our base case, we assumed that the probability of a “true case” being tested was *p*_test_=0.1 and the number of individuals tested for each true case was *n*_test_=0.5; we assumed a mean reporting delay between onset of infectiousness and testing results of 6.6 days (24). We then modelled scenarios in which the fraction of true cases detected and reported (as indicated by *p*_test_) and the testing capacity (i.e. number of individuals tested for every true cases, *n*_test_) either increased or decreased linearly between days 20 and 60 of the epidemic. To examine the impact of sudden changes to the probability of a true case being tested (e.g. associated with changes in testing criteria) and testing capacity (e.g. associated with new companies entering the market), we also explored scenarios in which *p*_test_ and *n*_test_ increased or decreased abruptly on day 40 (Table S1). Finally, we examined the effect of a two-fold increase or decrease in the average reporting delay.

### Estimation of R_0_ from simulated data

We estimated *R*_0_ from the growth rate of the epidemic, as described by Lipsitch et al (11):

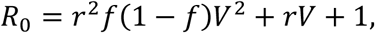

where *r* is the growth rate, *V* is the serial interval (also known at the generation interval), and *f* is the proportion of the serial interval spent in the latent period. The growth rate (*r*) was determined by fitting Poisson regression models to the cumulative number of “true cases” (*Y*_*d*_) and reported cases (*C*_*d*_) on days *d*=21 to 40. Thus, we implicitly assumed that cases occurring over the first 20 days of the epidemic are unlikely to have been recognized. We assumed that the serial interval is equal to the sum of the average latent period and the average infectious period (*V*=1/*ν* + 1/*γ*), and that the proportion of the serial interval spent in the latent period is *f*=1/(*Vν*). We estimated 95% confidence intervals (CIs) for our estimate of *R*_0_ by incorporating uncertainty in the estimated growth rate, but did not incorporate uncertainty in *f* or *V*.

### Estimation of R_t_ from simulated data

We estimated *R*_*t*_ using the Wallinga-Teunis method (7). This is a likelihood-based framework that infers the estimated number of secondary cases per case at each point in time based on information about the generation interval. First, the relative likelihood that case *i* was infected by case *j* is estimated based on the probability distribution of the difference in the time of symptom onset (*t*_*i*_ *– t*_*j*_), over the probability that case *i* was infected by any other case *k*:

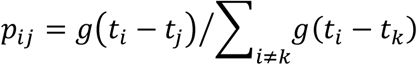

where *g*(*x*) is the probability distribution function for the generation interval. We assume all cases are equivalent in terms of infectiousness. Thus, to estimate *R*_*t*_ based on the number of “true” cases, this can be rewritten as:

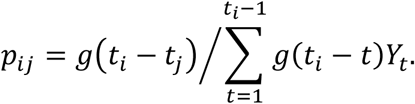

We do not allow for values of the generation interval less than or equal to zero; thus, we assume all cases were infected by a case that occurred on an earlier day. While this is not necessarily true when accounting for possible reporting delays, we make the same assumption when estimating *R*_*t*_ based on the number of reported cases. We assumed that the generation interval was gamma distributed with a mean of 6.5 days and coefficient of variation of 0.62, consistent with data from Flaxman et al (20):

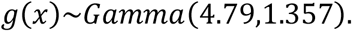

Second, the reproductive number for case *j* is estimated by calculated the weighted sum of *p*_*ij*_ over all cases *i*:

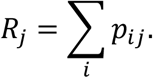

Again, assuming all cases are equally infectious, this becomes:

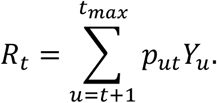

To estimate *R*_*t*_ based on the number of true cases, we set *t*_*max*_=70 and estimate *R*_*t*_ up to day 60. We correct for right-censoring of the case data by dividing our estimates of *R*_*t*_ by the cumulative distribution function of the generation interval (*G*(*x*)) for *t*_*max*_ *– t*:

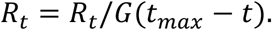

To explore the potential influence of the reporting delays, we estimate *R*_*t*_ based on the reported positive cases for *t*_*max*_=70 and *t*_*max*_=95.

### COVID-19 testing data for the United States

Daily data on the reported number of positive and negative tests for COVID-19 in the US and by state were downloaded from www.covidtracking.com on April 18, 2020 (12). The COVID Tracking Project data comes from state/district/territory public health authorities, and occasionally, from trusted news sources, official press conferences, or (rarely) social media updates from state public health authorities or governors [12]. All data sources are documented in a spreadsheet. We analyzed the data from March 4, 2020, onward, as this is the first date that negative tests for COVID-19 were consistently reported for the entire US.

Testing practices vary considerably between states, and some states changed their guidelines a few weeks into the outbreak, depending on test availability, outbreak intensity, and health systems infrastructure. We extracted information on COVID-19 testing criteria from each state’s website during the week of March 23 and again during the week of April 13. Data sources are documented in the SI Dataset.

For example, as of March 13, Louisiana recommended testing for people with fever, respiratory symptoms, and a negative influenza test, with priority given to the following categories:

- Hospitalized patients with a severe respiratory illness with no other known cause.
- Suspected outbreak of COVID-19 among associated individuals with recent onset of similar fever and lower respiratory symptoms.
- Recent fever and lower respiratory symptoms in a healthcare worker with direct contact to a laboratory-confirmed COVID-19 case.
- Suspected COVID-19 in a patient associated with a high-risk exposure setting such as a long-term care facility or a correctional facility.
- Suspected COVID-19 in a homeless patient.

However, as of April 15, anyone with fever and respiratory symptoms was eligible to be tested, regardless of influenza status, and pharmacists were given permission to order and administer COVID-19 tests for the duration of the public health emergency.

### Estimation of R_0_ from state-level testing data

To estimate *R*_0_ from data on the number of reported positive COVID-19 cases in the US and different states, we fitted Poisson regression models to the first three weeks of data (March 4 through March 24, 2020) to estimate the growth rate. For states that did not report any cases early on, the Poisson regression models were fitted to the data beginning the first day a positive case was reported through March 24. This is approximately one week after the national “15 Days to Slow the Spread” guidelines were announced (on March 16, 2020) (25). We assumed that the mean serial interval was 6.5 days and that the average latent period was 4.6 days, based on (20). We calculated 95% CIs for the *R*_0_ estimates based on uncertainty in the estimated growth rate, but did not account for uncertainty in the serial interval or latent period.

### Estimation of R_t_ from state-level testing data

Estimates of the time-varying reproductive number in each state were generated using the Wallinga-Teunis method described above. We again assumed that the generation interval was gamma distributed with a mean of 6.5 days and coefficient of variation of 0.62 (20). Estimates after April 5, 2020, are increasingly uncertain because <95% of secondary cases are expected to have occurred by the end of the available data (based on the cumulative distribution function of the generation interval). For later dates, we present both the uncorrected estimates and estimates corrected for right-censoring, as described above.

## Data Availability

All data used in this analysis are publicly available from https://covidtracking.com/

## Acknowledgements

This work was supported by the following grants from the National Institutes of Health/National Institute of Allergy and Infectious Diseases (NIH/NIAID): R01AI137093 (VEP, JLW, DMW), R01AI112970 (VEP), R01AI123208 (DMW), and R01AI146555 (NAM, TC).

## Supporting Information

**Table S1.** Scenarios for simulated changes in testing practices.

| Scenario | Fraction of true cases detected and reported ($p_{\text{test}}$) | Testing capacity, i.e. number of individuals tested for each true case ($n_{\text{test}}$) | Reporting delay distribution parameters |
| --- | --- | --- | --- |
| 1. Constant testing effort | 0.5 | 5 | $a=1.85, b=3.57$ |
| 2. Linear increase in testing probability | 0.05 on $d=20$ to 0.15 on $d=60$ in increments of 0.01 | 5 | $a=1.85, b=3.57$ |
| 3. Linear decrease in testing probability | 0.15 on $d=20$ to 0.05 on $d=60$ in increments of 0.01 | 5 | $a=1.85, b=3.57$ |
| 4. Linear increase in testing capacity | 0.5 | 0.2 on $d=20$ to 0.8 on $d=60$ in increments of 0.015 | $a=1.85, b=3.57$ |
| 5. Linear decrease in testing capacity | 0.5 | 0.8 on $d=20$ to 0.2 on $d=60$ in increments of 0.015 | $a=1.85, b=3.57$ |
| 6. Abrupt increase in testing probability | 0.05 for $d \leq 30$<br>0.15 for $d > 30$ | 5 | $a=1.85, b=3.57$ |
| 7. Abrupt decrease in testing probability | 0.15 for $d \leq 30$<br>0.05 for $d > 30$ | 5 | $a=1.85, b=3.57$ |
| 8. Abrupt increase in testing capacity | 0.5 | 0.2 for $d \leq 30$<br>0.8 for $d > 30$ | $a=1.85, b=3.57$ |
| 9. Abrupt decrease in testing capacity | 0.5 | 0.8 for $d \leq 30$<br>0.2 for $d > 30$ | $a=1.85, b=3.57$ |
| 10. Increase in reporting delay | 0.5 | 5 | $a=1.85$ for $d \leq 30$<br>$a=3.7$ for $d > 30$<br>$b=3.57$ |
| 11. Decrease in reporting delay | 0.5 | 5 | $a=1.85$ for $d \leq 30$<br>$a=0.925$ for $d > 30$<br>$b=3.57$ |

**Table S2.** Estimates of the basic reproductive number, *R*_0_, for different US states. Estimates are based on fitting the growth rate in the number of confirmed cases or total number of tests performed through March 24, 2020, using Poisson regression, and assume a fixed serial interval of 6.5 days. The 95% confidence intervals (CI) incorporate only uncertainty in the estimated growth rate.

| State abbrev. | $R_0$ (based on confirmed cases) | 95% CI (cases) lower bound | 95% CI (cases) upper bound | $R_0$ (based on number of tests) | 95% CI (tests) lower bound | 95% CI (tests) upper bound |
| --- | --- | --- | --- | --- | --- | --- |
| AK | 4.25 | 3.47 | 5.23 | 3.05 | 2.98 | 3.12 |
| AL | 3.58 | 3.33 | 3.87 | 7.17 | 6.95 | 7.40 |
| AR | 3.72 | 3.46 | 4.01 | 3.59 | 3.50 | 3.69 |
| AZ | 4.16 | 3.91 | 4.40 | 2.33 | 2.26 | 2.39 |
| CA | 2.69 | 2.65 | 2.74 | 2.58 | 2.57 | 2.60 |
| CO | 3.01 | 2.92 | 3.10 | 3.12 | 3.08 | 3.15 |
| CT | 4.72 | 4.49 | 4.95 | 5.04 | 4.96 | 5.12 |
| DC | 3.23 | 2.99 | 3.49 | 4.02 | 3.91 | 4.12 |
| DE | 3.37 | 3.01 | 3.75 | 2.04 | 1.92 | 2.16 |
| FL | 3.60 | 3.51 | 3.69 | 4.19 | 4.15 | 4.22 |
| GA | 3.53 | 3.43 | 3.63 | 2.89 | 2.82 | 2.95 |
| HI | 3.71 | 3.30 | 4.18 | 11.86 | 11.46 | 12.30 |
| IA | 2.72 | 2.50 | 2.93 | 4.41 | 4.31 | 4.50 |
| ID | 3.63 | 3.06 | 4.25 | 3.19 | 3.12 | 3.25 |
| IL | 4.05 | 3.96 | 4.15 | 3.60 | 3.57 | 3.63 |
| IN | 4.09 | 3.85 | 4.33 | 4.64 | 4.54 | 4.75 |
| KS | 3.51 | 3.19 | 3.84 | 3.34 | 3.25 | 3.43 |
| KY | 3.04 | 2.81 | 3.29 | 3.57 | 3.50 | 3.65 |
| LA | 4.03 | 3.92 | 4.15 | 5.49 | 5.42 | 5.56 |
| MA | 2.86 | 2.79 | 2.93 | 4.04 | 3.99 | 4.08 |
| MD | 3.43 | 3.28 | 3.59 | 1.98 | 1.92 | 2.05 |
| ME | 3.38 | 3.10 | 3.71 | 3.16 | 3.11 | 3.20 |
| MI | 2.62 | 2.60 | 2.64 | 2.98 | 2.96 | 3.00 |
| MN | 3.25 | 3.08 | 3.44 | 2.75 | 2.73 | 2.78 |
| MO | 5.17 | 4.74 | 5.63 | 2.36 | 2.28 | 2.46 |
| MS | 4.82 | 4.50 | 5.15 | 3.43 | 3.35 | 3.51 |
| MT | 3.20 | 2.74 | 3.74 | 3.40 | 3.33 | 3.47 |
| NC | 3.76 | 3.59 | 3.95 | 4.29 | 4.23 | 4.34 |
| ND | 3.79 | 3.08 | 4.50 | 3.83 | 3.74 | 3.92 |
| NE | 2.36 | 2.15 | 2.59 | 2.68 | 2.60 | 2.75 |
| NH | 2.96 | 2.73 | 3.21 | 2.99 | 2.93 | 3.04 |
| NJ | 4.74 | 4.66 | 4.83 | 5.75 | 5.66 | 5.83 |
| NM | 2.72 | 2.43 | 3.00 | 3.42 | 3.38 | 3.46 |
| NV | 3.57 | 3.37 | 3.77 | 3.91 | 3.86 | 3.97 |
| NY | 4.74 | 4.71 | 4.78 | 4.26 | 4.25 | 4.28 |
| OH | 4.12 | 3.92 | 4.32 | 2.88 | 2.79 | 2.97 |
| **OK** | 3.46 | 3.14 | 3.78 | 3.03 | 2.95 | 3.10 |
| **OR** | 2.63 | 2.50 | 2.77 | 3.01 | 2.97 | 3.04 |
| **PA** | 3.75 | 3.62 | 3.87 | 4.25 | 4.20 | 4.30 |
| **RI** | 2.91 | 2.70 | 3.11 | 2.73 | 2.67 | 2.78 |
| **SC** | 3.78 | 3.59 | 4.00 | 3.60 | 3.53 | 3.66 |
| **SD** | 1.92 | 1.60 | 2.24 | 2.50 | 2.44 | 2.56 |
| **TN** | 4.05 | 3.89 | 4.20 | 5.88 | 5.80 | 5.97 |
| **TX** | 3.12 | 2.99 | 3.24 | 3.56 | 3.51 | 3.61 |
| **UT** | 3.81 | 3.61 | 4.03 | 4.66 | 4.58 | 4.73 |
| **VA** | 3.07 | 2.91 | 3.22 | 3.45 | 3.41 | 3.50 |
| **VT** | 3.95 | 3.55 | 4.37 | 2.75 | 2.69 | 2.80 |
| **WA** | 2.15 | 2.13 | 2.18 | 2.59 | 2.58 | 2.60 |
| **WI** | 3.81 | 3.66 | 3.98 | 4.11 | 4.07 | 4.15 |
| **WV** | 4.47 | 2.79 | 6.22 | 3.89 | 3.74 | 4.05 |
| **WY** | 2.89 | 2.43 | 3.43 | 3.12 | 2.97 | 3.29 |
| **US** | **3.448** | **3.435** | **3.461** | **3.530** | **3.525** | **3.535** |

**Fig. S1.**
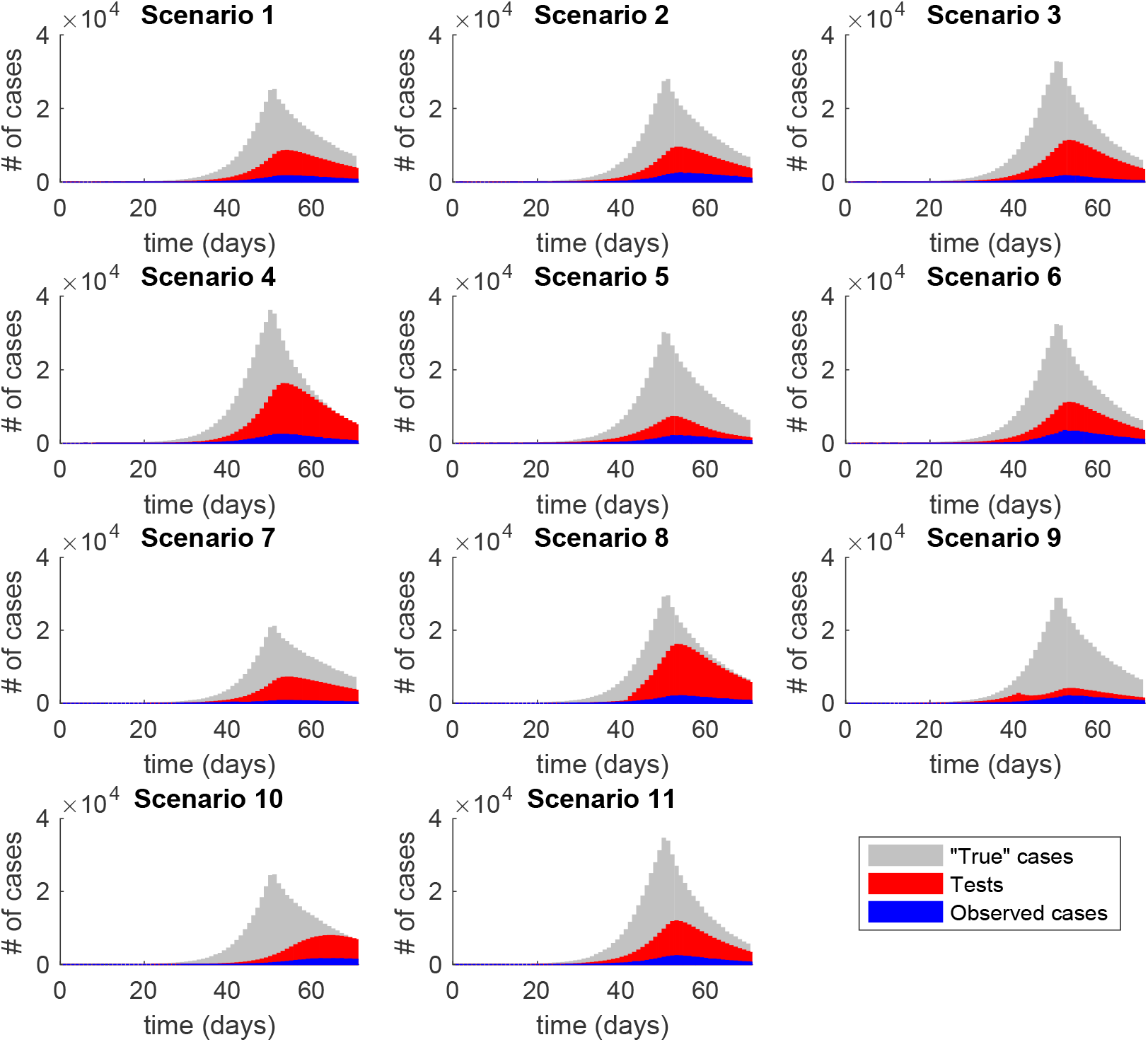
Simulated epidemics for different modelled scenarios exploring changes in testing practices. Number of incident infections (“true” cases, grey), individuals tested (red), and confirmed cases (positive tests, blue) predicted by a stochastic SEIR model for a population of 1 million. Modelled scenarios are described in Table 3.

**Fig. S2.**
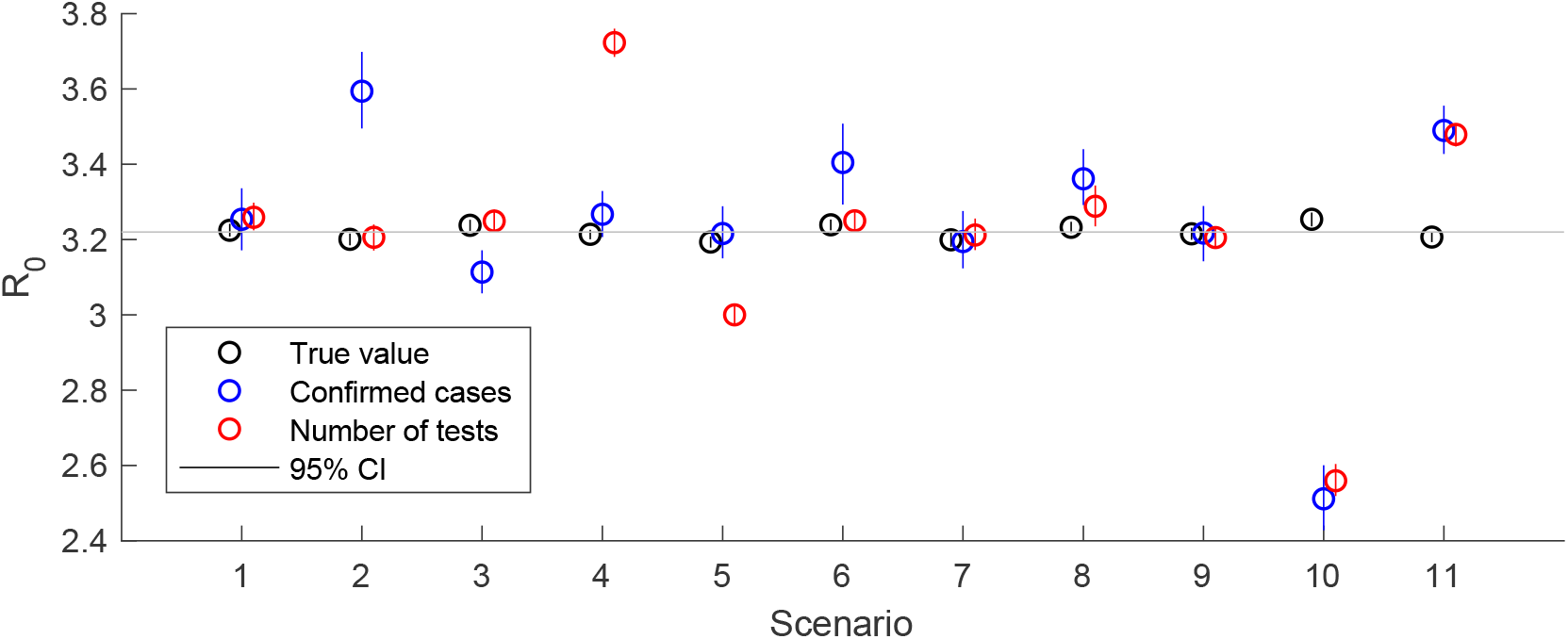
Estimates of the basic reproductive number, *R*_0_, based on simulated data. Estimated values of *R*_0_ based on the growth rate in the number of “true” cases (black), number of confirmed cases (blue), and number of tests performed (red) are plotted for the modelled scenarios described in Table 3. The mean value is represented by the open circle, vertical lines represent 95% confidence intervals (CIs) based on uncertainty in the growth rate. The grey horizontal line represents the mean value of the “true” *R*_0_ across all scenarios.

**Fig. S3.**
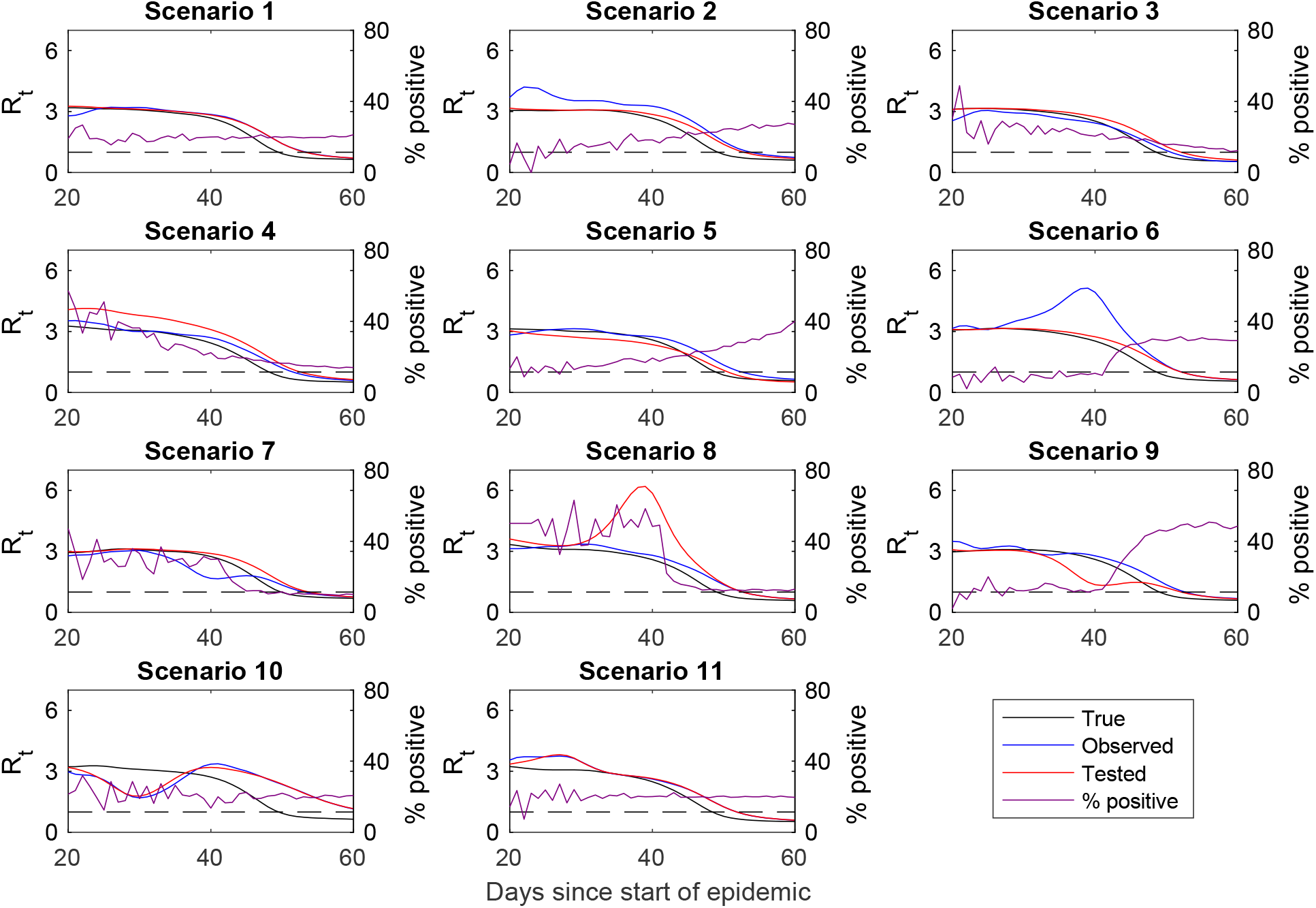
Estimates of the time-varying reproductive number, *R*_*t*_, based on simulated data. Values of *R*_*t*_ estimated by fitting to the number of “true” cases (black), confirmed cases (blue), and number of tests performed (red) are plotted along with the percent of positive tests (purple) for the modelled scenarios described in Table 3. The horizontal dashed line represents *R*_*t*_=1.

**Fig. S4.**
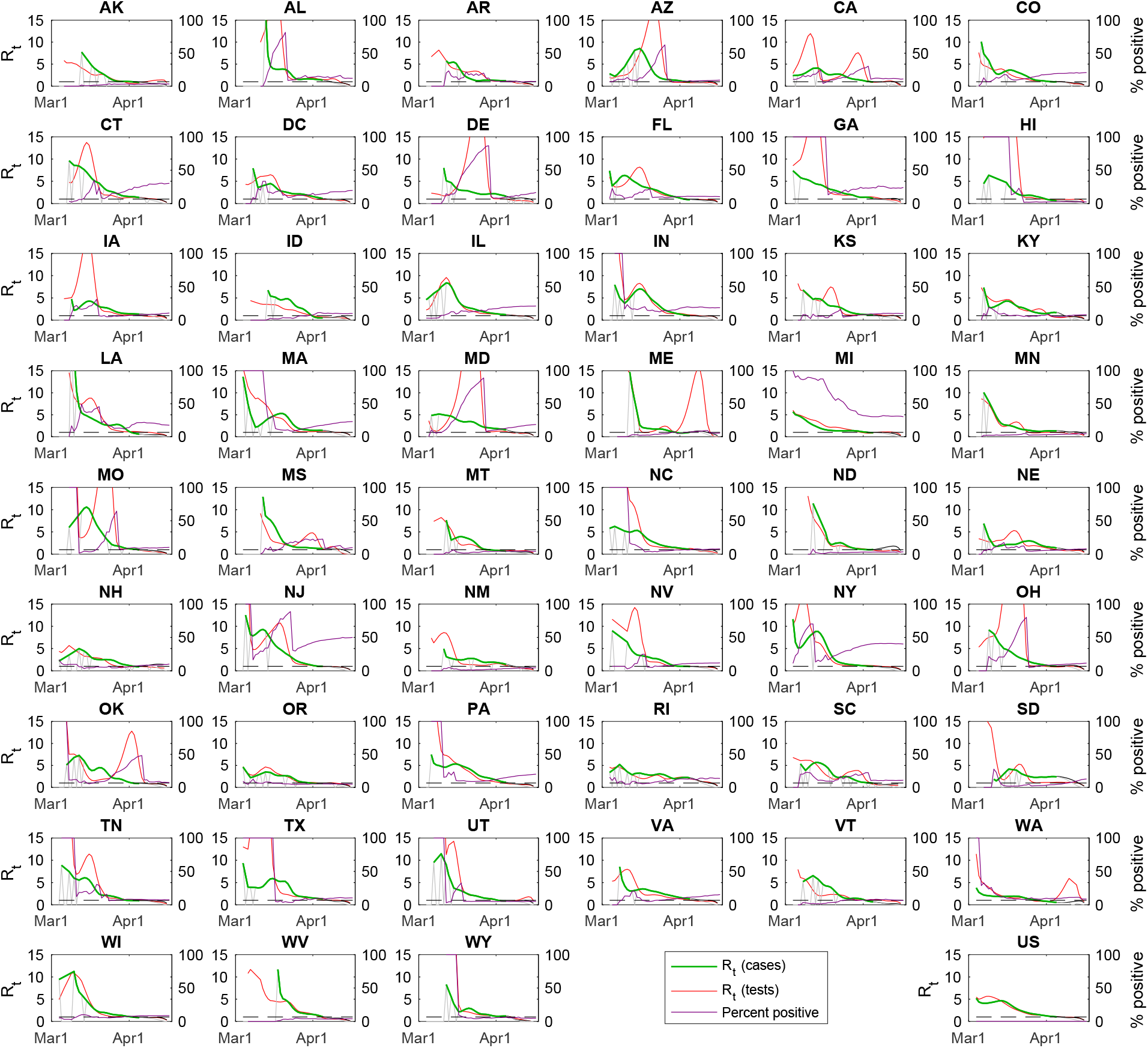
Estimated time-varying reproductive numbers for 50 US states and the District of Columbia. Estimates of the time-varying reproductive number, *R*_*t*_, based on the number of confirmed cases (green) and the total number of tests performed (red) are plotted for each state for March 4 to April 17, 2020, along with the percent of tests positive (purple). April 5 is the last day that the value of *R*_*t*_ can be reliably estimated; uncorrected (grey) and corrected (black) values of *R*_*t*_ are plotted for April 2 to April 16. Value of *R*_*t*_ based on the number of tests are corrected for right-censoring.

